# Co-designing a virtual reality based mindfulness application to address diabetes distress using Artificial Intelligence-informed Experience-Based Co-Design (AI-EBCD): a feasibility study

**DOI:** 10.64898/2026.03.10.26348062

**Authors:** Shraboni Ghosal, Mengying Zhang, Emma Stanmore, Jackie Sturt, Angeliki Bogosian, David Woodcock, Nicola Milne, Womba Mubita, Glenn Robert, Siobhan O’Connor

## Abstract

More than one third of adults with diabetes can experience diabetes distress due to the demands of daily self-care. As a cognitive therapy, mindfulness can alleviate diabetes distress but face-to-face programmes can be difficult to access and pay for, and apps lack personalisation and feedback. Virtual reality (VR) may support mindfulness practice, but no VR app tailored to people experiencing diabetes distress exists. We interviewed mindfulness practitioners and conducted co-design workshops (using focus groups, questionnaires, artistic methods, generative artificial intelligence tools and prioritization techniques) with adults with type 2 diabetes to gather perspectives on designing a VR mindfulness app. We analysed data using descriptive statistics and the framework approach. Most participants preferred a simple design and layout to use the virtual environment to practice mindfulness, with customisable design options and interactive features that were culturally appropriate. We identified new design features, functionality, and content that informed a software design specific documentation to build a prototype VR mindfulness app for people experiencing diabetes distress. Further research should include more diverse populations to elicit detailed specifications for software design and include safety features to minimise risk when using VR technologies to practice mindfulness.

## Introduction

Type 2 diabetes is a chronic metabolic disease that is a major global health issue estimated to affect up to 642 million people by 2040 ^1,2^. Individuals with type 2 diabetes are at a higher risk of developing complications such as cardiovascular disease, kidney damage, nerve damage and vision loss ^3^. Management of type 2 diabetes involves long-term lifestyle changes, frequent monitoring of blood sugar levels, along with medications to control blood sugar levels ^1^. However, many individuals suffer experience emotional burden and stress related to managing diabetes which includes feelings of being emotionally and physically overwhelmed by the daily demands of the disease ^4,5^. Approximately 36% of people with diabetes experience diabetes distress ^5^ which can negatively impact self-management, with poorer glycaemic control and increased risk of complications ^4,6^. This may lead to additional medical care and psychological interventions that increase healthcare costs ^5^ which are already significant, with the United States spending an estimated $237 billion on diabetes care in 2017 ^7^ and the United Kingdom (UK) spending on average £10 billion per year, around 10% of the entire budget of the National Health Service ^8^.

Treating diabetes distress requires interventions such as counselling, cognitive-behavioural therapy, mindfulness based interventions, and specialist diabetes education ^9^ which can improve glycaemic control, may help to prevent complications and contribute to savings long-term ^5^. Mindfulness is a third wave cognitive therapy, rooted in Buddhist traditions, that helps people stay focused in the present moment without judgments ^10^ by emphasising awareness of thoughts, feelings, bodily sensations, and the environment, without becoming overwhelmed or overly reactive ^11^. Mindfulness-based interventions have been used to manage diabetes distress, with promising results such as reduced emotional distress and improved quality of life ^12^. Mindfulness-Based Stress Reduction and Mindfulness-Based Cognitive Therapy are two programmes that can help individuals with diabetes manage distress, incorporating concepts of kindness and compassion, yoga practices, and mindfulness of body and breath and are also linked with for oneself and others ^13,14^. However, mindfulness can be difficult to adopt, with environmental and personal distractors often cited as challenges when practising. Face-to-face mindfulness offers a more personalised and deeper experience but can be less accessible, for example, for people living in remote areas or having busy schedules ^15^. The cost of in-person sessions led by instructors can be also prohibitive, especially for individuals with lower-incomes and the quality of mindfulness experience in these settings may also vary ^15^.

Mindfulness programmes have increasingly moved from traditional face-to-face settings to online methods with advances in digital technologies and growing demand. Online mindfulness programmes offer greater accessibility and convenience ^16^ and several mindfulness app such as Headspace, iMindfulness, and Mindfulness Daily have been rated as engaging and visually appealing ^17^. However, digital forms of mindfulness come with challenges such as issues with engagement, accountability, quality control, a lack of personalised feedback and guidance that in-person sessions provide ^16,17^. These can lead to more ad-hoc and less effective practice, especially for people with specific needs and the absence of an instructor or group can make people lose motivation ^18,19^. Virtual reality (VR) technology enables users to be fully immersed in virtual environments using a VR headset and hand controllers which may help overcome some of the limitations of face-to-face and digital mindfulness programmes. VR-based mindfulness applications can enhance user focus on the present moment by fully capturing their attention, leading to a deeper mindfulness effect compared to traditional methods ^20,21^. VR can also provide an immersive, interactive environment for practicing mindfulness by supporting the user to direct attention to the present moment within a tailored VR setting ^22,23^. However, VR has some drawbacks such as users experiencing cyber-sickness (i.e., dizziness, nausea) from prolonged periods of VR use which may affect the acceptability and efficacy of VR for mindfulness practice ^24^.

There is potential for VR mindfulness interventions to reduce stress and anxiety and improve overall well-being. Lower stress levels and improved emotional regulation were found in those who used VR to practice mindfulness compared to those who did not by enhancing emotional resilience and self-regulation ^24^ which may help manage diabetes distress. The adaptability of VR technology may also allow for the creation of personalised mindfulness experiences tailored to individual needs, such as those of people with type 2 diabetes, who face unique emotional burdens ^25^. In addition, VR apps can also be accessed from home, allow individuals to engage in mindfulness practices at their own convenience and pace, which may improve adherence and outcomes in managing diabetes distress ^26,27^. However, a recent review of VR apps for mindfulness showed no studies used a co-design approach to create the VR mindfulness experience with end users from beginning to end, with only existing VR apps or prototypes developed by research teams tested with them ^27^. Therefore, how a VR app should look and function to support people with type 2 diabetes to practice mindfulness to address diabetes distress is largely unknown, with previous research providing limited insights. Hence, our feasibility study aimed to understand the virtual experiences expected in a VR mindfulness app as this could offer an innovative approach to managing the emotional challenges associated with diabetes.

## Methods

We employed a multi-method approach over two phases of research via the collection, analysis of quantitative and qualitative data and used the Good Reporting of a Mixed Methods Study checklist for reporting purposes ^28^ (Supplementary File 1).

### Phase 1

We began by undertaking a commercial health app review of VR apps related to mindfulness and meditation, following clear methodological guidelines ^29^. We identified five such VR apps across the major app stores and utilised the Mobile App Rating Scale to assess their engagement, functionality, aesthetics, and information quality focusing that were available on VR app stores ^30^. This review helped us determine the availability and quality of existing VR-based mindfulness applications ^31^ which informed interviews with mindfulness experts and co-design workshops with people with type 2 diabetes. We then employed purposive sampling to recruit mindfulness practitioners over email, via contact information on the British Association of Mindfulness-based Approaches website (www.bamba.org.uk) and our professional networks, to take part in semi-structured interviews. We developed, piloted and refined an interview guide to explore the key concepts and practices of mindfulness and the perceptions of expert practitioners on what virtual experiences they would like in a VR app for mindfulness. This included showing the participants images and video clips of existing VR mindfulness apps (i.e., Headspace XR, Hoame, Innerworld, Maloka and TRIPP) identified from our commercial health app review ^31^ to stimulate in-depth discussion. Key characteristics of a VR-based mindfulness intervention (e.g., timing, frequency, duration) were also discussed to help establish when, how often, and for how long it should be used (Supplementary File 2). In addition, we used a short demographic questionnaire to capture key participant characteristics (Supplementary File 3). One researcher (SG) conducted the individual semi-structured interviews with mindfulness practitioners (n=9) in March 2024 over Zoom. Each interview lasted approximately 60 minutes, and automatic transcription was enabled to generate a transcript. These interviews along with the commercial health app review were used to inform phases 2 and 3 of the study.

### Phase 2

Participatory design is a popular way to create digital health technologies with end users to ensure a high-quality digital health product or service is developed that meets the needs of those who will use it ^32,33^. This process can include several phases from brainstorming the initial problem and potential solutions, to creating prototypes, and developing and testing software and/or hardware with end users ^34^. By employing a range of participatory methods at each stage ranging from interviews, focus groups, and observation, to personas, storyboarding, and prototyping, a better quality and more usable digital health tool is likely to be developed which could increase user engagement and improve health outcomes long-term ^32,35^. Hence, we adopted a well-developed and extensively used participatory design framework called Experience-Based Co-Design (EBCD) to support phases 2 and 3 of the study ^36,37^. We adapted EBCD by incorporating artistic practices and generative artificial intelligence (AI) tools (i.e., audio, video, image and music AI generators) into the design process to enable a richer understanding of what people with type 2 diabetes wanted from a VR mindfulness app. Generative AI tools can create a range of digital media quickly by responding to text-based prompts from human users ^38^. Therefore, they represent a novel way for people to explore their creativity and express their preferences for audio-visual content that can be incorporated into digital technologies ^39^. We coined this new participatory approach Artificial Intelligence-informed Experience-Based Co-Design (AI-EBCD). We adopted a purposive sampling approach to recruit adults with type 2 diabetes to a series of co-design workshops. We posted advertisements on several social media platform and distributed through diabetes charities and online university forums to recruit participants to the five co-design workshops (Table 1).

**Table 1.** Workshops conducted as part of the AI-EBCD approach.

| Workshop 1 | Workshop 2 | Workshop 3 | Workshop 4 | Workshop 5 |
| --- | --- | --- | --- | --- |
| <u>Participants</u>: adult with type 2 diabetes (n=14)<br><u>Aim</u>: to discuss diabetes distress, learn about and discuss mindfulness practice<br><u>Method</u>: online focus group via Zoom<br><u>Tools</u>: emotional mapping to discuss diabetes distress, and mindfulness training. (question guide, observation sheet, and a demographic questionnaire, supplementary files 3 and 4)<br><u>Timeframe</u>: May 2024 (90 minutes)<br><u>Facilitators</u>: SG and AB | <u>Participants</u>: adult with type 2 diabetes (n=12)<br><u>Aim</u>: to experience and evaluate existing commercial VR apps for mindfulness (i.e., Headspace XR, Hoame, Innerworld, Maloka and TRIPP)<sup>31</sup><br><u>Method</u>: online workshop via Zoom<br><u>Tools</u>: video and audio clips of commercial VR mindfulness apps, Mobile App Rating Scale<sup>30</sup><br><u>Timeframe</u>: May 2024 (90 minutes)<br><u>Facilitators</u>: SG | <u>Participants</u>: adult with type 2 diabetes (n=1)<br><u>Aim</u>: to use artistic methods to create a personalised mindfulness experience<br><u>Method</u>: drawing/sketching or other artistic methods<br><u>Tools</u>: creative materials were posted to participants who were also encouraged to utilise any additional resources at their disposal to create artistic outputs<br><u>Timeframe</u>: June 2024 (2 weeks)<br><u>Facilitators</u>: SG and SOC | <u>Participants</u>: adult with type 2 diabetes (n=12)<br><u>Aim</u>: to use a suite of generative AI (GenAI) tools to create digital content for a VR mindfulness app<br><u>Method</u>: individual think aloud interviews while using GenAI tools<br><u>Tools</u>: think aloud protocols (supplementary file 5), licenses for four GenAI tools including image, audio, video, and music generators (i.e., Picsart, MURF AI, Soundraw, and Pictory) with all digital content created downloaded, short presentation on the ethical and socio-legal issues with GenAI to educate participants<br><u>Timeframe</u>: June 2024 (60 minutes each)<br><u>Facilitators</u>: SG | <u>Participants</u>: adult with type 2 diabetes (n=7)<br><u>Aim</u>: to prioritise key elements for a VR mindfulness app that were identified during the previous four co-design workshops<br><u>Method</u>: online focus group via Zoom<br><u>Tools</u>: nominal group technique<sup>40</sup> used to individually and collectively score and rank the elements allowing equal participation, ‘round robin’ technique within the group discussion to resolve disagreements<br><u>Timeframe</u>: June 2024 (60 minutes)<br><u>Facilitators</u>: SG |

The results of the prioritisation exercise in workshop five enabled us to draft a software design document to support the development of a prototype VR mindfulness app in phase 3. Generative AI platforms are a novel co-design tool that can support people to express their imagination and design preferences using different forms of digital media ^39,41^. However, due to copyright concerns the GenAI outputs from workshop four were not used directly to develop any of the design features or digital content for the VR mindfulness app but helped inform workshop five and the final software design specification document.

### Data analysis

We analysed data after each phase of the study to enable the preliminary findings to inform future phases of data collection and analysis (SG, SO). We analysed all qualitative data using the framework approach to enable the identification of themes and sub-themes ^42^. This involved 5-stages: (i) familiarisation (ii) constructing a thematic framework (iii) indexing and sorting (iv) data summary and display, and (v) mapping and interpretation. N-Vivo QSR 14.0 was used to support the analytical process. First, familiarisation took place, we listened to audio files from interviews and focus groups and read transcripts several times, checking for accuracy and completeness, and noting key ideas and recurring concepts. We reviewed drawings from the third workshop and the artistic data (i.e., images, audio, video and music) generated during the fourth co-design workshop and from these, we created an initial set of codes, so they emerged inductively from the data to address the research aims. We compared and contrasted the codes to structure them into two preliminary thematic frameworks (addressing each research question) and cross-checked these before the next round of analysis. We then applied the descriptive frameworks to a number of transcripts to refine, and group related codes into index or overarching categories reflecting emerging similarities, differences and relationships across participant groups (i.e., mindfulness practitioners, adults with type 2 diabetes). They were then systematically applied across the remaining transcripts to identify patterns and associations in the data while developing the thematic frameworks further. Next, we undertook charting to rearrange categories into thematic headings and subheadings, grouping abstracted and synthesised cases (i.e., a distilled summary of participants views or experiences) under relevant subjects to enable cross-comparison between or within cases. In some of these cases data saturation was reached. All quantitative data from the co-design workshops were analysed using descriptive statistics on Microsoft Excel.

### Rigour and reflexivity

Rigour was enhanced through periodic peer debriefing to ensure the research process and interpretation of data were credible, with samples of analytical coding checked, helping minimise researcher bias ^43^. Clear descriptions were provided of all methods to enhance dependability, with video and audio-recordings compared against transcripts to ensure accuracy, and qualitative quotes for themes provided to support the findings. To increase transferability and applicability of the study’s findings to other areas, the context was described in detail, with the strengths and limitations of the research highlighted.

### Ethics

Ethical approvals were received from [blinded for peer-review] University Research Ethics Committee for phases 1 and 2 (Ref: 2024-18262-32710) and phase 3 (Ref: 2024-21170-37093) of the study.

## Results

### Participant characteristics

The nine mindfulness practitioners who were interviewed in phase 1 were aged between 46-58 years (M=52, SD=5.4), with 5 being women and 4 men, all of White-British origin. They were all well-educated, with one having a doctorate degree, four had Master’s degrees, two had Bachelor’s degrees and one’s level of education was unknown (Table 1). Most (78%) were experienced having 3 to over 15 years professional experience as a mindfulness practitioner. The thirteen adults with type 2 diabetes who took part in the series of co-design workshops in phase 2 and the two participants from the user evaluation in phase 3 were aged between 24-54 years (M=32.6, SD=8.2), with 10 being men and 3 women, most being White British origin (n=11, 85%) and the origin of two participants unknown. Most were well-educated with 2 having Master’s degrees, 7 had Bachelor’s degrees, one had college qualifications, one was an apprentice and the level of education of two participants was unknown. All thirteen self-reported a diagnosis of type 2 diabetes from about 1 to 25 years (Table 2).

**Table 2.** Sociodemographic characteristics of participants.

| Sociodemographic characteristics | Mindfulness Practitioners |  | Adults with type 2 diabetes |  |
| --- | --- | --- | --- | --- |
|  | N | % | N | % |
| **Age** |  |  |  |  |
| 18-29 | 0 | 0 | 6 | 46 |
| 30-39 | 0 | 0 | 5 | 39 |
| 40-49 | 3 | 33 | 1 | 8 |
| 50-59 | 5 | 56 | 1 | 8 |
| 60+ | 0 | 0 | 0 | 0 |
| Unknown | 1 | 11 | 0 | 0 |
| **Gender** |  |  |  |  |
| Female | 5 | 56 | 3 | 23 |
| Male | 4 | 44 | 10 | 77 |
| **Race/ethnicity** |  |  |  |  |
| White-British | 9 | 100 | 2 | 15 |
| Black/Black-British | 0 | 0 | 5 | 38 |
| Mixed origin | 0 | 0 | 6 | 46 |
| **Education** |  |  |  |  |
| A Levels/BTEC/Apprenticeship | 0 | 0 | 2 | 15 |
| Bachelor's degree | 2 | 22 | 7 | 54 |
| Masters | 4 | 44 | 2 | 15 |
| PhD | 1 | 11 | 0 | 0 |
| Unknown | 2 | 22 | 2 | 15 |
| **Years of mindfulness experience / type 2 diabetes diagnosis** |  |  |  |  |
| 0 - 1 year | 1 | 11 | 6 | 46 |
| 2-5 years | 2 | 22 | 4 | 30 |
| 6-10 years | 2 | 22 | 1 | 8 |
| 11-15 years | 1 | 11 | 0 | 0 |
| 15+ years | 3 | 33 | 2 | 15 |
| **Employment** |  |  |  |  |
| Full-time | 2 | 22 | 6 | 46 |
| Part-time | 7 | 78 | 6 | 46 |
| Voluntary | 0 | 0 | 1 | 8 |

Two themes emerged relating to the expectations of a mindfulness experience within a virtual reality environment which were: 1) design and features of a VR environment for mindfulness and 2) mindfulness VR content.

### Theme 1. Design and features of a VR environment for mindfulness

The layout and visual appeal of a VR mindfulness app was discussed by numerous participants, with most preferring clear and simple navigation and the use of large easy to read fonts to make the VR app quick and easy to set up and use for different types of people with diabetes distress (Table 3). A relaxing colour palette with natural imagery was also of importance to some participants (Figure 3). Participants also requested features in the VR app that made it simple and uncomplicated to use such as a free trial period to encourage mindfulness practice, with some practitioners believing this would make the app beneficial as an adjunct to face-to-face classes. The ability to customise the VR app was a function many participants requested as they wanted to tweak the design (e.g., colour palette, graphics, icons), tailor some of the app features (e.g., avatars, notifications/reminders, user profiles, personal targets, language, privacy and security settings) and personalise the mindfulness content (e.g., audio, images, video) to create a VR experience that suited their preferences and needs. Interactive features were also high on participants priority list, as they valued aspects that encouraged VR app use to support mindfulness practice. This included functions such as a tracker for mood, games and quizzes as incentives, the ability to provide feedback, and connecting and sharing progress with app ‘friends’. One mindfulness practitioner suggested a virtual forum within the VR app where users could discuss the benefits and limitations of their practice with others, while another highlighted the importance of including a safety trigger in the VR app for anyone who had negative experiences so they could get support to manage diabetes distress.

**Figure 3.**
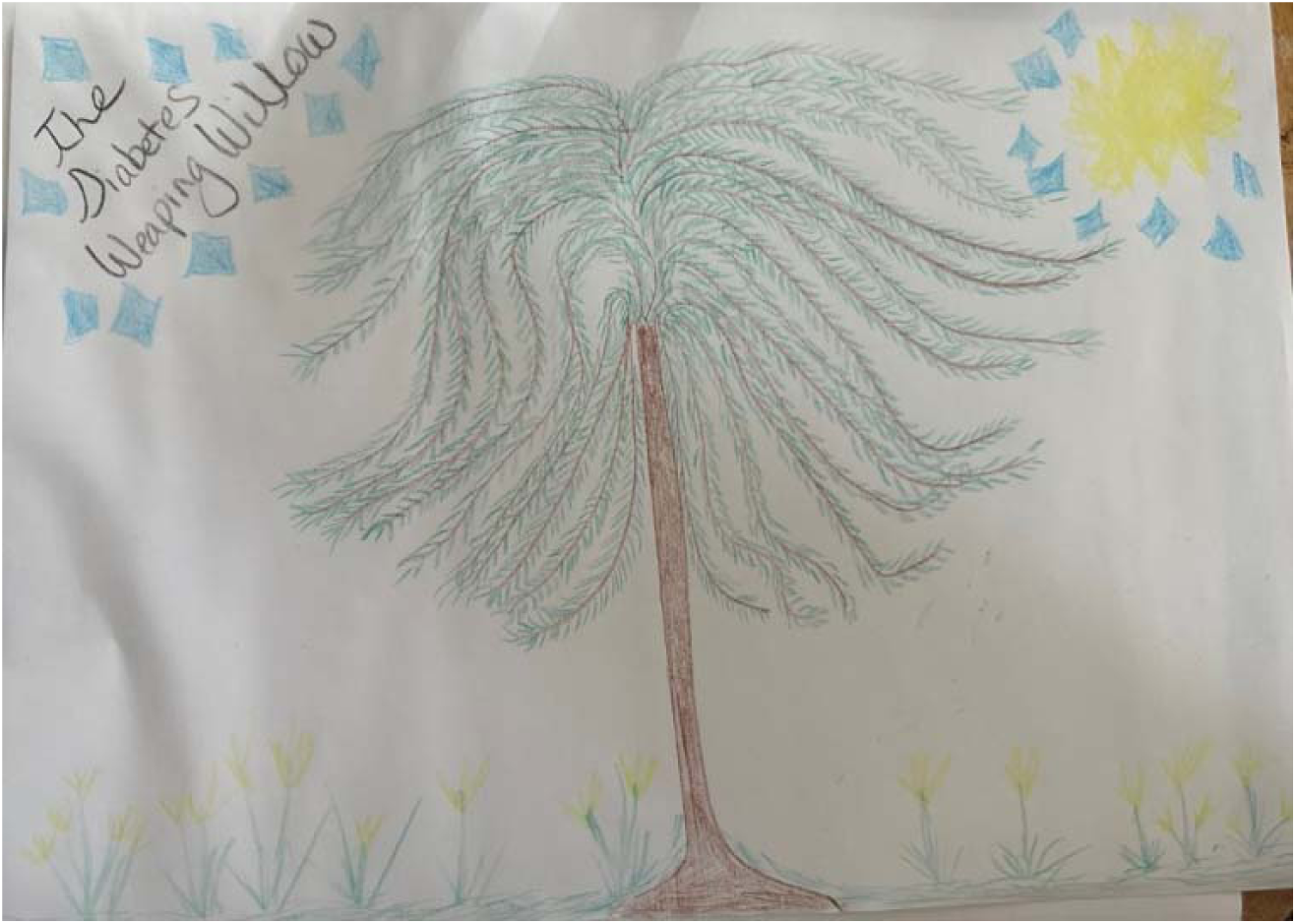
Artistic sketch of a personalised mindfulness experience (Person with T2D, Workshop 3)

**Table 3.** Participant quotes to support theme one.

|  |
| --- |
| *“What would I think if I was just, you know, designing something like that? I think I would say keep it really simple. Don't overcomplicate it because otherwise then the brain starts getting into into doing mode doesn't it? It starts, it starts. You know, you'd have to. If you've got lots going on which they did seem to be in those apps to be honest, there was a lot going on you're already in thinking mode aren't you so you're already not really being mindful because you're doing far too much thinking whereas mindfulness is about being more in that state of being so it would It would really need to complement that”*<br>(Mindfulness Practitioner Interview 6, Female) |
| *“There is a need for....the very essence of it would be a clear navigation.....we can actually set up font that are actually customised, I think that will be better”*<br>(Person with T2D, Workshop 5) |
| *“Where you can make it really easy and just go on your phone and find a meditation. If that helps to stimulate your interest in mindfulness and helps you to embed. A regular practice. Then I would encourage it”*<br>(Mindfulness Practitioner Interview 9, Male) |
| *“it would be good to be able to customise the background a little bit, maybe a bit like the avatar, to be able to customise that, so it can resemble you or something completely different to give you that break from your illness”*<br>(Person with T2D, Workshop 5) |
| *“But if there was anything that could help draw you back into it so that you are more likely. Especially if you're not on a course anymore, you know, or in between the weeks of a course that would something that would draw you back in see. Doing a bit of practice and giving you a bit of positive reinforcement and a bit of advice and guidance and I just think Yeah, I think that's where it's, I think that's would be really helpful”*<br>(Mindfulness Practitioner Interview 2, Female) |
| *“I loved what we did in designing our own videos and songs and tracks. It would be lovely to be able to have that artistic input in some some form, whether it's to create our own avatar or a theme music, something where we have some sort of either functionality”*<br>(Person with T2D, Workshop 5) |
| *“I think interactivity is good, you know, in the form of enforcement, feedback, quizzes,* |
| *games, anything that you can do to hook people into coming back to something, being excited about it, wanting to do it. I just think you have to make sure it's rooted in mindfulness"*<br>(Mindfulness Practitioner Interview 2, Female) |
| *"if I had a reminder that would be good, set up a reminder and to do it regularly like...once once a day would be good. I think if I did it once a day. I would get more out of it than just doing it when I kind of feel, you know I should do it... Yeah, that would be good."*<br>(Person with T2D, Phase 3 Interview, Female) |
| *"It could, you know, the way, you know, I talked about the 3-minute breathing space or a body scan or a breathing practice or a movement practice. I mean if it had those in the VR in the app, you could people could then go do it on their own or they could maybe join a group that was doing that practice and then if there was a conversation after that practice about the practice and then they could also go, I would have thought, and joined groups not practicing, but talking about whether they've managed to practice or not and how that had felt or not if the you know what stops them coming you know practicing or you know they could be, you know, more talking about mindfulness in that way"*<br>(Mindfulness Practitioner Interview 2, Female) |
| *"I'd want to know that there was some kind of recognition in there or that they have a almost like a safe word or a button to press if they need. You know, if they're struggling with it because otherwise if they just take it off and you're leaving them with no support and that could be problematic"*<br>(Mindfulness Practitioner Interview 1, Female) |

### Theme 2. Mindfulness VR content

Many participants wanted to see a variety of audio narrations supporting mindfulness practice in a VR environment especially ones which were culturally appropriate as they felt this would be essential core content, although a few practitioners raised the point that naturals sounds are often relied on in face-to-face mindfulness sessions to stimulate authentic self-awareness and self-reflection which VR may not be able to replicate (Table 4). Some did consider listening to music as a form of mindfulness which could be added to a VR app but only where it was not a distraction. Adults with type 2 diabetes used the GenAI tool to create calm and peaceful soundscapes, with a slow to regular tempo, to accompany guided mindfulness narrations and/or visualisations.

**Table 4.**
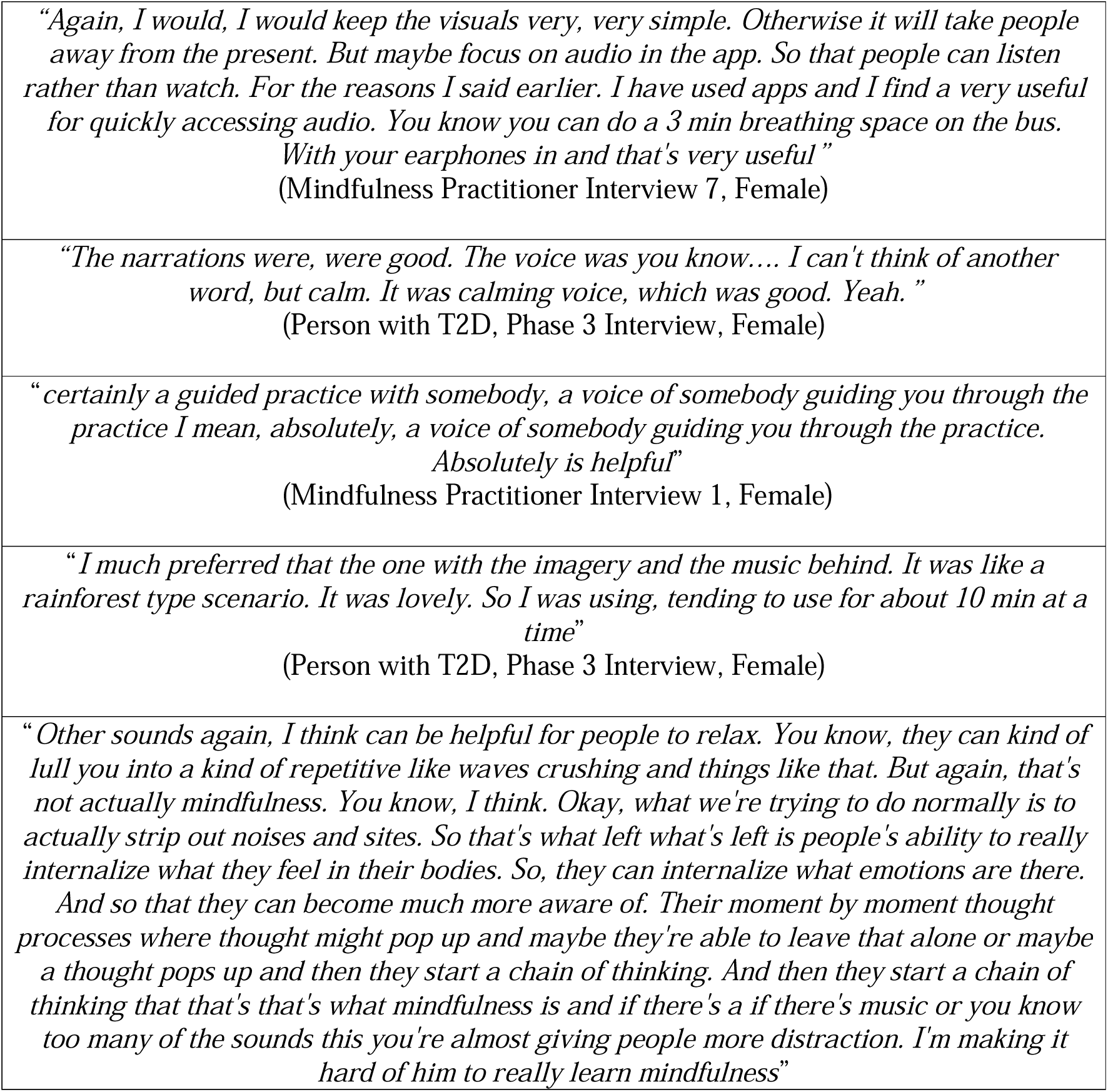
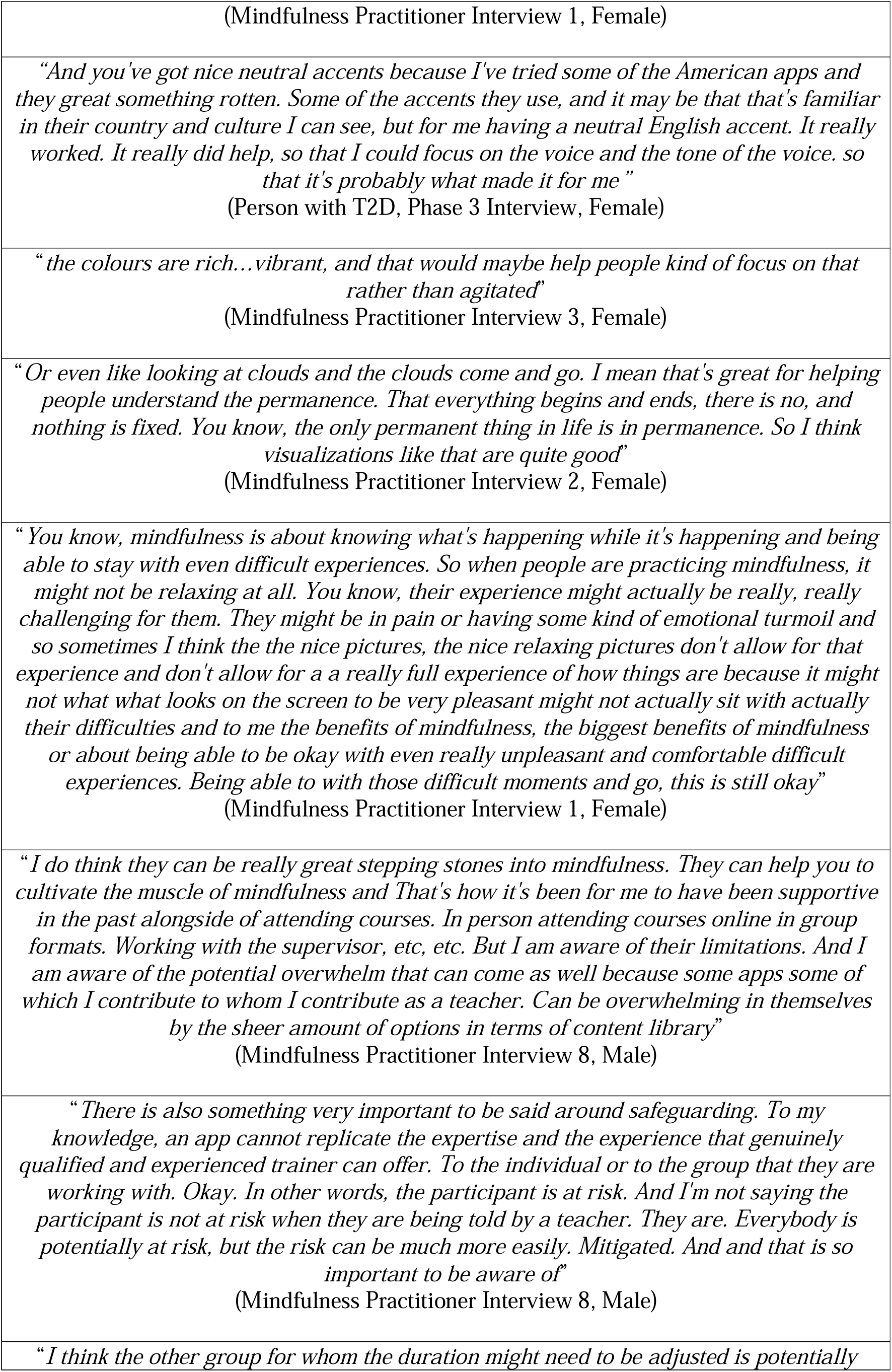
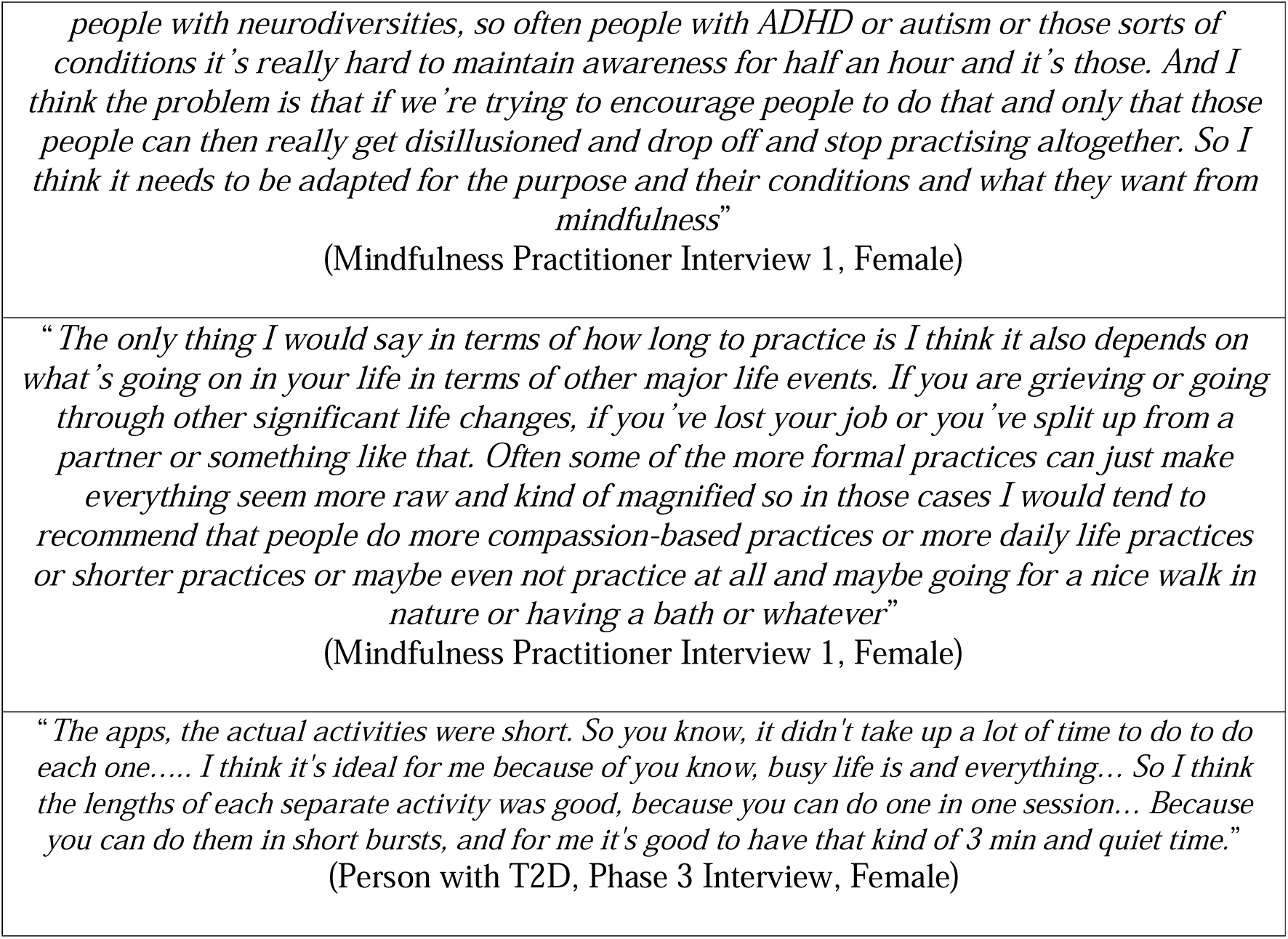
Participant quotes to support theme two.

Participants had mixed views about using visualisations to support VR-based mindfulness practice, with some seeing them as something positive that people could focus on to help escape agitation or anxiety. High quality visualisations created by adults with T2D using GenAI tools largely centred on nature-based imagery such as peaceful outdoor landscapes some which included humans in meditative poses (Figures 4 and 5). However, several mindfulness practitioners felt visualisations could be distracting and take away from mindfulness practice particularly if someone was in pain or emotional turmoil as their feelings would not align with the VR content.

**Figure 4.**
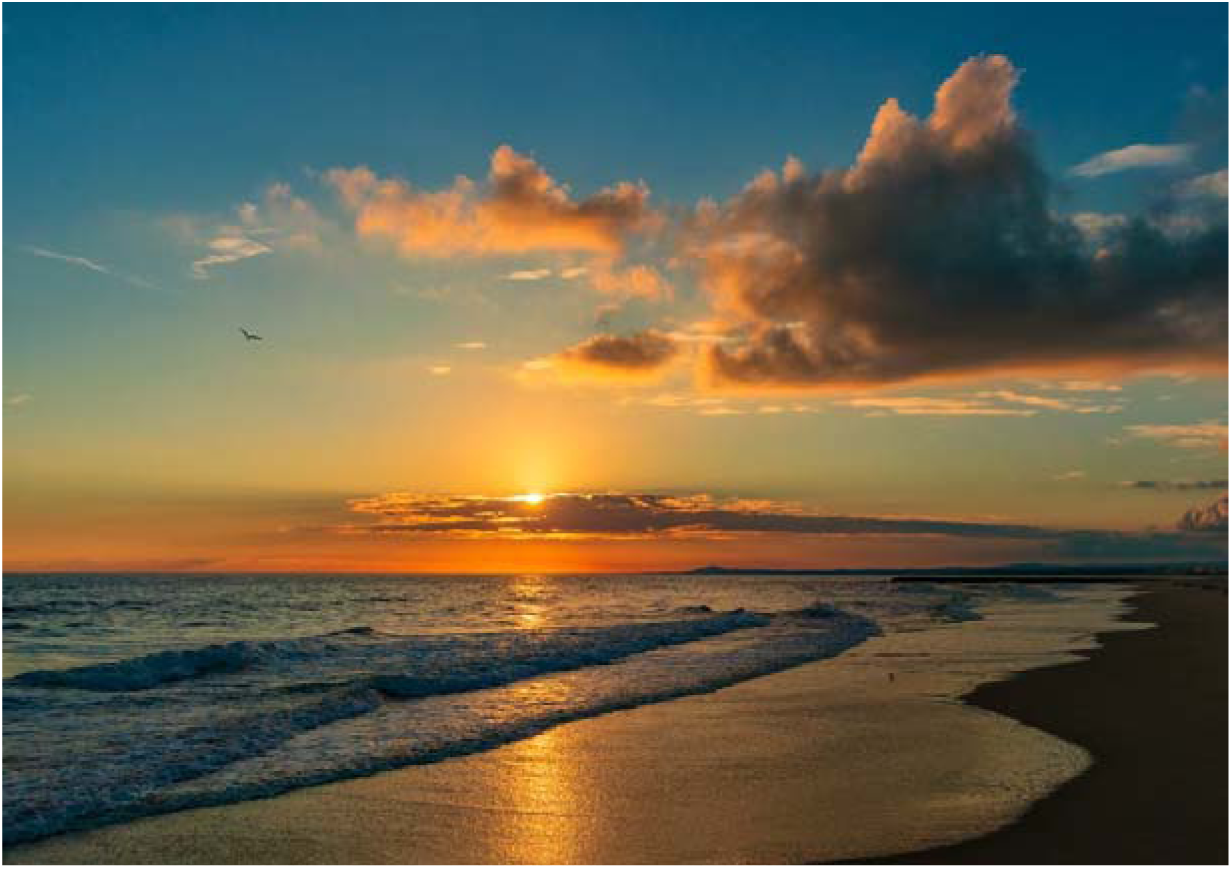
Similar image to that created via a GenAI tool (Person with T2D, Workshop 4) Victoria Yurch: https://www.pexels.com/photo/clouds-over-sea-shore-at-sunset-15790159/

**Figure 5.**
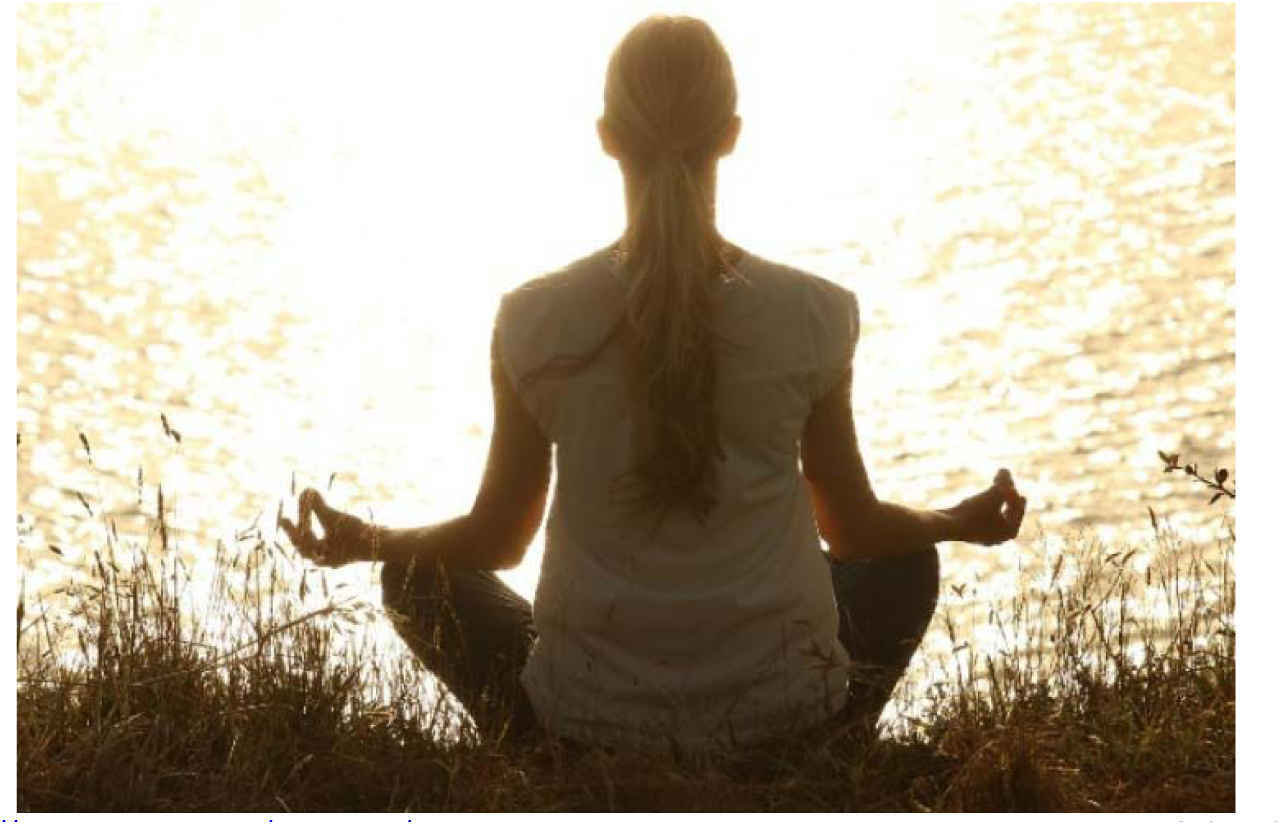
Similar image to that created via a GenAI tool (Person with T2D, Workshop 4) Pexels: https://pixabay.com/photos/meditate-woman-yoga-zen-meditating-1851165/

Other types of content adults with T2D wanted from a VR app for mindfulness included inspirational stories or quotes from other diabetes users, transcripts of different mindfulness practices, and educational content about diabetes and mindfulness, with some emphasising the need for regular content updates to keep people using the VR app. However, one practitioner warned of information overload in providing too much mindfulness and other content to users and stressed the importance of striking a balance between attending in person classes (individual or group based) alongside using a mindfulness app (Table 3). Other practitioners highlighted the potential risk of an individual practising mindfulness via a VR app as there would be limited support mechanisms in place if they had a distressing experience while engaging with the content. Bearing that in mind, some practitioners said they would recommend apps for practicing mindfulness as the scientific evidence base for them is growing. The duration of mindfulness practice was also discussed with a range of views from shorter sessions that were only a few minutes to longer practices that were half an hour or more in length depending on the individuals’ personal circumstances.

## Discussion

This study gathered the perspectives of mindfulness practitioners and adults with type 2 diabetes on the key elements they would like to see a VR app for mindfulness to help alleviate diabetes distress. Most wanted a simple layout and design where they could navigate around the virtual environment easily to access and engage with a range of mindfulness practices. Customisation was also sought after by many participants who wanted to be able to tailor various design elements and functionality in the VR app to suit their needs, while interactive features such as mood tackers and games were also requested to help sustain use of the app long-term. Cultural adaptation was also thought to be important so the virtual mindfulness content could showcase audio, images, and video content that suited users’ diverse needs, although many participants had a preference for natural landscapes and soothing sounds. Regular content updates were also a priority to make the app appealing to users. However, some mindfulness practitioners were sceptical of using visualisations as they felt it would be a distraction to mindfulness practice. The safety of users was another concern as people may recall negative or distressing experiences when practising mindfulness which requires professional support.

The growing body of literature on using VR for mindfulness practice across different patient and general populations have reported some similar results. For instance, Seabrook et al. ^24^ who trailed a VR mindfulness app in a general population found the visual and auditory elements of the virtual experience helped participants focus on the present moment enabling them practice mindfulness, while a review of commercial VR apps for mindfulness found several used natural environments, guided mindfulness narration, and had some customisable features to engage users ^31^. However, in many research studies participants were provided with an existing VR mindfulness app or one developed by the research team to test its effectiveness in improving health outcomes and were not involved in co-design, with limited to no exploration of the design features, function and content of a VR mindfulness app preferred by end users ^27^. Saying that some of these studies reported using natural scenarios overlaid with mindfulness narration as a means of delivering virtual mindfulness practice ^20,27,44^ which aligns with our findings. Furthermore, a scoping review of VR based mindfulness interventions for chronic pain found a number of usability issues such as cybersickness, the need for technical support, and more personalised design where users could curate their own virtual worlds to help them engage in mindfulness practice ^23^.

Only two other studies adopted a co-design approach to create a new VR app for mindfulness. Hugh-Jones et al.^45^ worked with adolescents who had a history of mental health difficulties and mental health and education professionals to develop a solution with them to manage teenage stress. However, priority setting for the app was completed by professionals and only a prototype was tested with the young people, two of whom reviewed the mindfulness audio but not the VR environment, excluding them from in-depth involvement in the design of the VR mindfulness app. The user testing did reveal adolescents wanted both day and night environments, a choice of VR environments to suit their mood, and better mindfulness narration with introductions and spaces to practice mindfulness in silence. Kelly et al.^46^ combined top down and user centred activities to design a VR mindfulness app for general users. However, much of the exploratory design work was undertaken by the research team guided by evidence and experts, using video footage of a real forest combined with voiceovers of mindfulness practice, with nine end users only involved at the prototype testing phase. The users highlighted the importance of good sound quality, with variations in the duration of mindfulness practice, and alternative positions for practice such as lying down as useful features of a VR app. Finally, O’Gara et al.^47^ also co-designed a virtual reality intervention with young people undergoing cancer treatment, but this focused more on relaxation and compassion training than mindfulness. Similar to our study, they adopted an EBCD approach without using artistic or generative AI methods during the design process and reported high-quality VR imagery was valued by participants and real virtual environments over animated ones.

The integration of VR into mindfulness practice to support people with diabetes distress presents unique opportunities and challenges. Future research would benefit from larger and more diverse sample sizes that include a range of genders, ages, ethnicities and experiences with mindfulness practice to elicit detailed design requirements for a VR mindfulness app as these are largely missing from the literature. As people with type 2 diabetes lead complex lives and can struggle with healthy eating, being physically active, and managing medication among other issues^1^, a more tailored VR mindfulness experience aligned with these daily self-care tasks could be beneficial for helping them manage diabetes distress. While reviewing the evidence on VR for mindfulness and speaking to expert practitioners are useful ways to elicit design ideas for a VR mindfulness app ^41^, the lived experience of those with diabetes is critical to include but a broader range of approaches to co-design could support more nuanced design and lead to a better-quality VR app for mindfulness for this population. An online methods toolkit on digital health co-design (https://www.digitalhealthcodesign.co.uk/) could inform future research as it details the mindsets, methods, tools and theories that can be used in the participatory design of digital health interventions such as VR apps. This would enable a deeper exploration of the types of mindfulness content that is needed, additional interactive features users want in a VR mindfulness app, and how much customisation would benefit engagement with the app and mindfulness practice. The safety aspects of using VR technologies, whether low or high-fidelity, for mindfulness also warrant more attention. Prior research report VR related issues such as cybersickness, eye strain, and the need for technical support ^27^ but not a negative mindfulness experience within a VR environment and whether professional or other support would be needed to manage this.

### Strengths and limitations

This study employed a robust co-design framework adapted to include generative AI tools in the design process and gathered diverse data from participants with different perspectives on mindfulness and virtual reality. International reporting guidelines were also used alongside the framework approach to analyse data with several techniques employed to enhance qualitative rigour. However, the small sample of mindfulness experts and adults with type 2 diabetes in our feasibility study limits the findings, and those with type 2 diabetes were not experienced in practising mindfulness which may have influenced their design needs and perspectives on a VR app for mindfulness. Hence, the results should be interpreted with caution as they may not be generalisable to diverse diabetes populations in the UK and internationally, where diabetes care, mindfulness practice, and the use of digital mental health interventions can vary considerably.

## Conclusion

The use of VR to support mindfulness practice is an emerging field particularly in diabetes care. This feasibility study incorporated evidence synthesis, the guidance of expert practitioners, and adapted a widely used co-design framework integrating artistic methods and generative AI tools, to design a VR mindfulness app with adults with type 2 diabetes to manage diabetes distress. Although this study is preliminary, its findings show the design features, functionality and content adults with type 2 diabetes want in a VR app for mindfulness. Future research should include more diverse participants to delve further into these to understand how to tailor a VR app to an individuals’ needs and incorporate safety features to minimise any risk using VR technology to practice mindfulness. Once a well-designed VR mindfulness intervention is available for people with type 2 diabetes, then clinical trials can be conducted to examine its efficacy in improving physical and mental health outcomes. While these types of digital mental health interventions are relatively new in the field of diabetes, research in other chronic diseases indicates they could have value for those who need more support in managing diabetes distress.

## Author contributions

SO and NM conceptualised the study. SO, ES and NM secured funding. SO, ES, AB, JS, DW, GR and WM supported the study design and methodology. SG, MZ and SOC collected the data. SO and ES analysed the data. SO prepared the manuscript and all authors critically reviewed and approved the final manuscript.

## Acknowledgements

We would also like to thank the mindfulness experts and people with type 2 diabetes for their voluntary participation in this study. Although the development of the prototype VR mindfulness app was not reported in this study, we would like to thank Andrew Jerriosn and Andrew Rowlye from Research IT at the University of Manchester who developed the VR app using Unity (https://unity.com/) and C# scripting language, and GitHub (https://github.com) to manage the software development process. Finally, we would like to thank the PACT app team (https://pact-app.com/) for providing us with mindfulness recordings we used in this study.

## Data availability statement

Data not available due to ethical restrictions

## Abbreviations

AI: Artificial Intelligence
AI-EBCD: Artificial Intelligence-informed Experience-Based Co-Design
EBCD: Experience-Based Co-Design
GRAMMS: Good Reporting of a Mixed Methods Study
UK: United Kingdom
VR: Virtual Reality

